# A geospatial machine learning prediction of arsenic distribution in the groundwater of Murshidabad district, West Bengal, India: spatio-temporal pattern and human health risk

**DOI:** 10.1101/2022.05.21.22275403

**Authors:** Bibhash Nath, Antara Das, Tarit Roychowdhury, Wenge Ni-Meister, Mohammad Mahmudur Rahman

## Abstract

Arsenic (As) contamination of groundwater in parts of South and Southeast Asia is a public health disaster. Millions of people living in these regions could be chronically exposed to drinking water with As concentrations above the World Health Organization’s provisional guideline of 10 µg/L. Recent field investigations have shown that the distribution of groundwater As in many shallow aquifers in India and Bangladesh is evolving rapidly due to massive irrigation pumping. This study compares a decade-old dataset of As concentration measurements in groundwater with a dataset of recent measurements using geospatial machine learning techniques. We observed that the probability of As concentrations >10 µg/L was much greater in the regions between two major rivers than in the regions close to the Ganges River on the eastern border of the study area, where As concentrations >10 µg/L had been measured prior to 2005. The greater likelihood that As is present away from the river channel and is found instead in the interfluvial regions could be attributed to the transport and flushing of aquifer As due to intense groundwater pumping for agriculture. We estimated that about 2.8 million people could be chronically exposed to As concentrations >10 μg/L. This high population-level exposure to elevated As concentrations could be reduced through targeted well-testing campaigns, promoting well-switching, provisions for safe water access, and developing plans for raising public awareness. Policymakers could use the ternary hazard map to target high-risk localities for priority house connections of piped water supply schemes to help reduce human suffering.

**Key points:**

- A high-resolution predictive analysis was conducted using geospatial machine learning techniques to identify human suffering.
- A comparison of decadal arsenic measurements and a machine learning prediction suggests a shift in hotspot location.
- Groundwater in a region between two major rivers was found to be unsafe for agricultural and drinking purposes.

**Plain language summary:** We conducted a high-resolution predictive analysis using geospatial machine learning algorithms to identify the extent and hotspot location of arsenic (As) contamination in the Murshidabad district of West Bengal, India. The predictive analysis identified an area between two adjacent major rivers in which the probability of As concentrations >10 μg/L in groundwater is significantly greater than in other areas. There is a shift in As hotspot location from the regions near the river toward the regions between the two adjacent rivers, possibly due to intense groundwater pumping for agriculture. We estimated that about 1.6 million people could be at high-risk from drinking water contaminated by As concentrations >10 μg/L. Policymakers could use the hazard map and the analysis of treated piped drinking water networks to provide access to targeted safe water wells for affected households.

## 1. Introduction

Groundwater from both confined and unconfined aquifers in the Ganges River basin was, for a long time, considered to be the safest water resource for human consumption. Due to a rapid increase in the demand for groundwater for irrigation and drinking, millions of tube wells were drilled indiscriminately in the alluvial plains of the Ganges River during the 1970s. This widespread utilization of groundwater without thoroughly testing and chemically analyzing its quality has resulted in the arsenic (As) crisis that currently affects the population of almost the entire Ganges River basin (Chakraborty et al., 2002, 2009; Bhattacharya et al., 2003; Nath et al., 2009; Ravenscroft et al., 2009). In the Ganges River basin, it was estimated that over 50 million people could be drinking water with As concentrations above the World Health Organization’s provisional guideline of 10 µg/L (Ahmed et al., 2006).

The discovery of As and its clinical manifestation were first reported in the West Bengal state of India in 1984 (Garai et al., 1984). Following the discovery of As in groundwater in West Bengal, numerous researchers have studied the As contamination and its health consequences for consumers (McArthur et al., 2004; Rahman et al., 2005; Charlet et al., 2007; Datta et al., 2011; Das et al., 2021). At the same time, sampling of well-water As over an extended area in West Bengal was carried out by the late Prof. Dipankar Chakraborti and his group at the School of Environmental Studies (SOES) at Jadavpur University. Their dataset documents the distribution of As over large contiguous areas of West Bengal before 2005 (Samanta et al., 1999; Chakraborti et al., 2009). The results indicated the widespread and heterogeneous distribution of As in alluvial aquifers. Most of the high-As concentrations were limited to the shallow alluvial aquifers (<300 feet) along and east of the Hooghly/Bhagirathi (hereafter called Bhagirathi) River, a distributary of the Ganges River (Chakraborti et al., 2009). For instance, the greater proportion of As in three concentration ranges in the district of Murshidabad in West Bengal are confined to the region located east of the Bhagirathi River (Rahman et al., 2005). The affected areas are predominantly low-lying, topographically flat, and located within the flood plains of networks of rivers draining the Himalayas (Nath et al., 2005; Fendorf et al., 2010).

In West Bengal, the groundwater occurs within the highly permeable alluvial formations deposited by the Ganges River. Irrigation pumping from shallow aquifers during dry seasons (January to May) for Boro rice cultivation draws a significant amount of water from extended geographical areas, causing a large cone of depression to form and the creation of a preferential flow path, allowing the penetration of As or reactive dissolved organic carbon to release As (van Geen et al., 2011; Erban et al., 2014; Khan et al., 2019). Moreover, deeper aquifers, mostly low in As, are extensively pumped to provide municipal water supply, causing the water level to drop significantly within a small area (Sahu et al., 2013). The unprecedented groundwater development was in response to the needs for securing pathogen-free drinking water and attaining self-sufficiency in food production (Harvey et al., 2005). Massive pumping of groundwater for irrigation and municipal water supply has thus substantially changed the natural groundwater flow system (Mukherjee et al., 2011) and has caused a rapid decline in the water table (15 cm/yr, Panda et al., 2022), which further impacts the long-term geochemical stability of As in the aquifers. Prior studies have shown that As release into the alluvial aquifers of the Ganges River is primarily driven by the microbially-mediated reductive dissolution of Fe-oxyhydroxides coupled to oxidation of sedimentary organic matter (Nickson et al., 1998; McArthur et al., 2001; Harvey et al., 2002; Smedley and Kinniburgh, 2002; Islam et al., 2004; Nath et al., 2008).

Excessive pumping of an aquifer can be a dominant factor controlling the natural groundwater flow system because it induces a lateral and vertical hydraulic gradient at regional and local scales (Michael and Voss, 2009). This practice could introduce contaminants to fresh, potable groundwater resources, resulting in devastating effects. Excessive pumping can also influence changes to redox conditions along a flow path. A recharge of anoxic waters to otherwise oxidized zones could cause a reduction of Fe(III) hydroxides, releasing sorbed As into groundwater. In contrast, the recharge of oxic water could reverse the ongoing redox reaction. The introduction of dissolved constituents, such as reactive organic carbon, from the surface to the aquifer could facilitate microbially-mediated redox reactions such as Fe-oxyhydroxide reduction, which leads to As release from sediments into groundwater (Datta et al., 2011). The changes in pH in groundwater as an effect of pumping could create a geochemical condition suitable for adsorption/desorption reactions to happen, resulting in the destabilization of As in the aquifer. Groundwater residence time and redox kinetics influence the stability of As in the aquifer. The enhanced flow velocity of groundwater could quickly disperse the dissolved As and other contaminants and redistribute them to the aquifer from where it was once released. Therefore, it is essential from a public health perspective to determine the current vulnerability of aquifers in response to massive groundwater pumping compared with a decade-old dataset on groundwater As concentrations to help identify hotspot locations for future intervention.

The spatial heterogeneity of As in the aquifers in the study area makes it difficult to predict its occurrences and understand long-term aquifer As stability. The cause of this heterogeneity is still a matter of debate despite intensive research of the studied region over the past three decades. It was proposed that the spatial heterogeneity of As in the aquifer of the Ganges River basin is strongly dependent on the redox chemistry, the source of organic matter, and the presence of clay lenses and palaeosols, i.e., the Pleistocene weathered land surfaces (van Geen et al., 2003; McArthur et al., 2004, 2011; Metral et al., 2008). A geospatial machine learning algorithm was developed to predict how As is distributed in the study area and determine the location of the highest vulnerability. A workflow was developed by recording the statistical relationship between predictor variables (representing climate, surface and sub-surface soil properties, and topography) and groundwater As concentrations. Machine learning models are powerful and have a great advantage over other conventional linear models because they can account for a non-linear relation between predictors and response variables (Ryo and Rillig, 2017).

Among machine learning algorithms, the random forest algorithm has been frequently used to model the spatial pattern of the probability of As exceedance in groundwater. This model also has been used to estimate the population at risk of exposure to elevated As and other environmental contaminants at regional and local levels (Podgorski et al., 2018; Podgorski and Berg, 2020; Tan et al., 2020; Mukherjee et al., 2021; Nath et al., 2022). These studies have illustrated the association between As occurrence and environmental predictors representing climatic conditions, soil characteristics, and topography. The associations between environmental predictor variables and As concentrations have been highlighted in previously published articles, such as how higher fluvisols’ probability could be associated with As occurrences and how the presence of surface clay could impact the spatial heterogeneity of As in groundwater (Nath et al., 2005; Donselaar et al., 2017; Wu et al., 2020). However, the scale of prior modeling studies was much greater (≥1 km), which could account for the small-scale spatial variation of As in groundwater (Winkel et al., 2008; Podgorski and Berg, 2020; Tan et al., 2021). Therefore, our main goals in this study were to predict As occurrences at a higher resolution (250 m) to capture spatial variability at habitation-scale, compare that variability to decadal-scale variability, and identify hotspot locations (and any shifts in hotspot location) in Murshidabad district—one of the most As contamination-affected districts of West Bengal, India (Chakraborty et al., 2009). This finer resolution study would produce a model with greater certainty and enable the effective implementation of targeted mitigation measures for greater public health benefits.

## 2. Study area

The study area, Murshidabad district, is located approximately 200 km north of Kolkata, the capital city of the West Bengal state of India (Fig. 1). The area is divided into two parts by the Bhagirathi River, a distributary of the Ganges River. The Ganges River borders the districts in the north and east. The district has a population of approximately 8.2 million people and covers an area of 5,550 km^2^. The climate is tropical, i.e., with wet and dry periods, and has an average monsoonal precipitation of 1,400 mm. We selected this study area because of two striking characteristics: the low proportion of high-As (>10 µg/L) concentrations in tube wells to the west of the Bhagirathi River, which reflects the presence of older Pleistocene aquifers (Chakraborty et al., 2009; Datta et al., 2011), and the proportion of high-As (>10 µg/L) concentrations vs. low-As (≤10 µg/L) concentrations to the east of the Bhagirathi River. Thus, this study area will enable us to understand whether there is any shift in the occurrence of As hotspots in response to the widespread use of groundwater for agricultural activity (Haque, 2015; Das et al., 2021).

**Figure 1.**
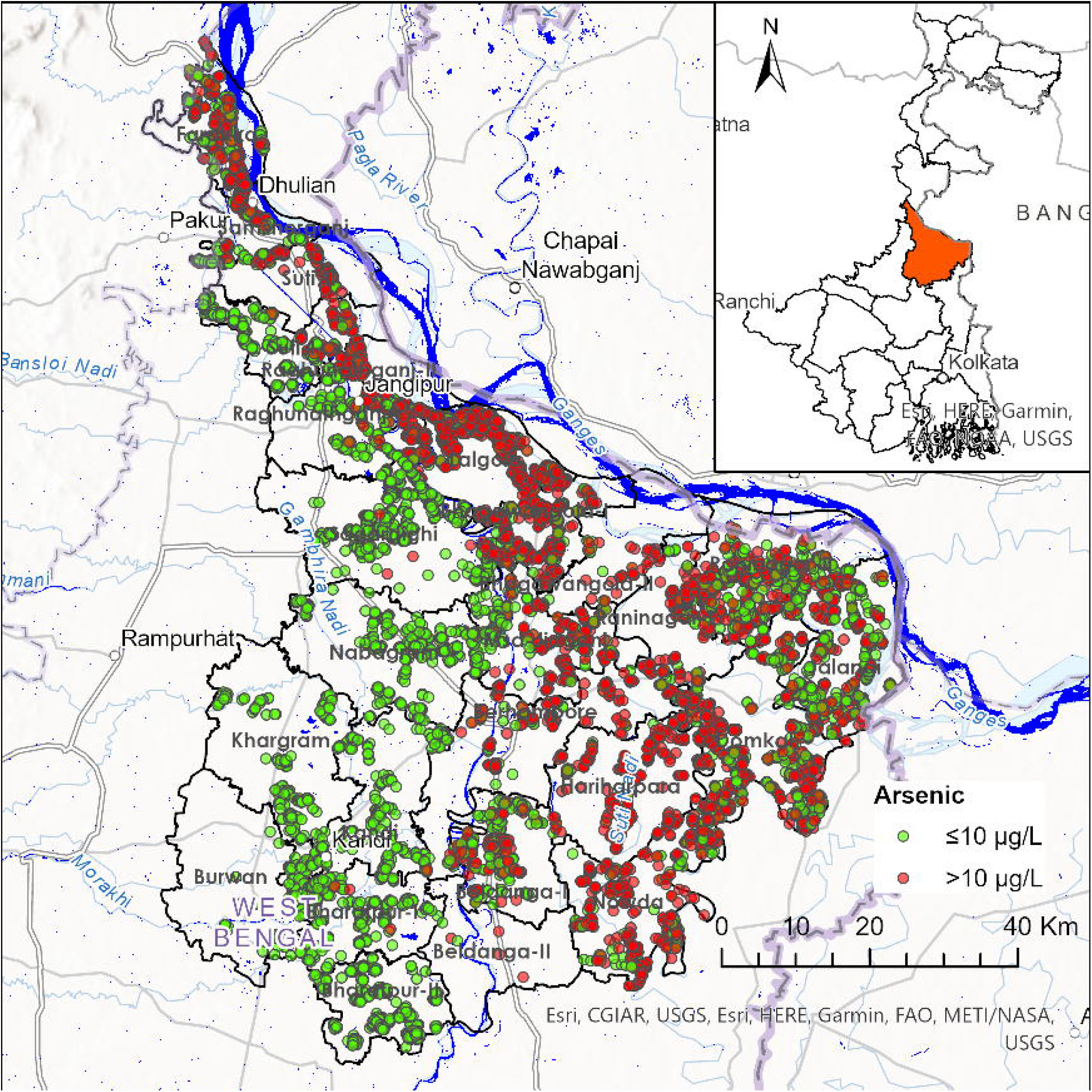
The map shows the location of the study area. The location (n=15,335) of tube wells measured for As concentrations are shown. Two major rivers, the Ganges and the Bhagirathi, and seasonal water areas (available for >6 months) are shown in dark blue (Pekel et al., 2016). The administrative boundaries demarcating 26 blocks (i.e., sub-districts) are also shown. The map in the inset shows India’s West Bengal state (with the capital city of Kolkata shown).

## 3. Methodology

### 3.1 Random forest model

The random forest model was carried out in the R environment (RStudio 2021.09.0) using the Caret package to develop a predictive relationship between environmental predictors and the occurrences of As concentrations >10 µg/L in groundwater (Kuhn, 2008). The random forest model is an ensemble of the decision tree, which is widely used in modeling the probability of As exceedance in groundwater based on linkages with geo-environmental predictors. The training of the model was done by the “bagging” method (i.e., a random selection of data with replacement). The random forest algorithm grows numerous decision trees on training data and gets predictions from each tree. The final, best prediction was based on majority voting. The majority voting method enables the model to produce more accurate predictions and helps reduce overfitting (Breiman, 2001). The random forest model is useful when several predictors are related. The use of predictors is restricted, and those predictors are randomly chosen during the development of a forest of decision trees (Podgorski et al., 2020), thereby allowing the model to produce an outcome with greater accuracy.

### 3.2 Arsenic in groundwater

We compiled As measurements of groundwater from tube wells in the Murshidabad district of West Bengal collected by Dipankar Chakraborti’s research group (Rahman et al., 2005). These datasets were compiled prior to 2005, and though they did not contain geolocations, they did have records of the block (sub-district)-level distribution of As in groundwater. For the predictive model, we compiled published data (from 2015 and later) of groundwater As concentrations with geolocation. These data are freely available for download from Public Health Engineering Directorate (PHED), West Bengal. The compiled datasets were cleaned to make sure the locations were correct based on other ancillary information recorded during the data collection, such as location within the requisite block boundary. We only used tube wells with depths ≤300 feet to make sure that our predictor variables could influence geochemical processes (Connolly et al., 2020). The cleaned dataset was grid-averaged at 250 m spatial resolution, and we calculated each grid cell’s geometric mean of As concentration. The grid-averaged As concentrations were then converted to a binary variable: 0 for As concentrations ≤10 µg/L and 1 for As concentrations >10 µg/L. As concentrations were converted to a binary variable to minimize the analytical uncertainty of the compiled data and to understand whether a particular location is in compliance with drinking water regulations.

### 3.3 Predictor variables

We used 26 predictor variables for model development (Table 1). These variables could be categorized as representing climatic conditions, soil characteristics, and topography. The selected predictor variables were directly or indirectly related to As occurrence in groundwater (Podgorski and Berg, 2020). The influence of these predictors was also checked using statistical relationships with the proportion of As concentrations >10 µg/L in groundwater. The statistical relationships were established by binning the predictor variables into 14 bins following the Sturges rule, keeping in mind that each bin contains an equal number of observations (Sturges, 1926). Mean concentrations in each bin were plotted against the percentage of As concentrations >10 µg/L. A Pearson correlation coefficient (R) and p-values were also checked for statistical significance.

**Table 1.** Spatially continuous predictor data sources were used in the development of a random forest model. The spatial resolution is provided. At the equator, 30” and 7.5” are approximately equal to 1 km and 250 m, respectively.

| **Predictors** | **Resolution** |
| --- | --- |
| Precipitation (Fick and Hijmans, 2017) | 30'' |
| Temperature (Fick and Hijmans, 2017) | 30'' |
| Potential evapotranspiration (PET) (Trabucco and Zomer, 2018) | 30'' |
| Aridity (Trabucco and Zomer, 2018) | 30'' |
| Topsoil organic carbon (Poggio et al., 2021) | 7.5'' |
| Subsoil organic carbon (Poggio et al., 2021) | 7.5'' |
| Topsoil sand (Poggio et al., 2021) | 7.5'' |
| Subsoil sand (Poggio et al., 2021) | 7.5'' |
| Topsoil silt (Poggio et al., 2021) | 7.5'' |
| Subsoil silt (Poggio et al., 2021) | 7.5'' |
| Topsoil clay (Poggio et al., 2021) | 7.5'' |
| Subsoil clay (Poggio et al., 2021) | 7.5'' |
| Topsoil cation exchange capacity (Poggio et al., 2021) | 7.5'' |
| Subsoil cation exchange capacity (Poggio et al., 2021) | 7.5'' |
| Topsoil nitrogen (Poggio et al., 2021) | 7.5'' |
| Subsoil nitrogen (Poggio et al., 2021) | 7.5'' |
| Topsoil pH (Poggio et al., 2021) | 7.5'' |
| Subsoil pH (Poggio et al., 2021) | 7.5'' |
| Topsoil bulk density (Poggio et al., 2021) | 7.5'' |
| Subsoil bulk density (Poggio et al., 2021) | 7.5'' |
| Fluvisols (Poggio et al., 2021) | 7.5'' |
| Gleysols (Poggio et al., 2021) | 7.5'' |
| Topsoil coarse fragments (Poggio et al., 2021) | 7.5'' |
| Subsoil coarse fragments (Poggio et al., 2021) | 7.5'' |
| Elevation (Hengl, 2018) | 7.5'' |
| Topographic wetness index (Hengl, 2018) | 15'' |

### 3.4 Model development and validation

The modeling was done at 250 m spatial resolution. The development of a high-resolution model enables a fine-scale understanding of the occurrence of As and whether rural inhabitants have access to safe water within 250 m. For the model development, the predictor’s information was collected in each grid location after converting the grids to a point geometry.

The data were divided into training (70%) and testing (30%) subsets. The ratio of high and low As concentrations was preserved through stratified sampling. The final selection of the predictors was based on recursive feature elimination, and it was conducted until the model did not suffer in accuracy or kappa values. The trained model was then used to predict the probability of As concentrations >10 µg/L for all study areas.

The scores of mean decrease in Gini and mean decrease in accuracy were computed to evaluate the relative importance of the predictor variables (Podgorski et al., 2018; Nath et al., 2022). More important variables have higher mean decrease accuracy and mean decrease Gini scores. Misclassification analysis is done by determining the model’s accuracy, sensitivity, and specificity (Mukherjee et al., 2021). A cutoff value distinguishes high and low As hazard areas. The cutoff value was chosen at the point where the model’s sensitivity and specificity intersect. The area under the receiver operating characteristic (ROC) curve was prepared to evaluate the predictive power of the model. The standard deviation of the classification outcome of all trees was computed to analyze model uncertainty. The lower standard deviation values determine the greater predictive power of the model.

### 3.5 Estimation of the total risk area and exposed population

The total at-risk area was estimated based on the probability cutoff values of the modeled prediction. The at-risk area was calculated by counting the number of pixels above and below the probability cutoff values in each block. Likewise, the total at-risk population was calculated using each pixel’s population density above probability cutoff values. These data were summarized for each block. The population density tile of 1 km spatial resolution for 2020 was collected from the WorldPop website (www.worldpop.org).

A ternary hazard map was prepared to estimate populations living in different risk areas of Murshidabad district. The ternary risk areas were defined as: high (probability ≥0.7), moderate (probability 0.56–0.7), and low (probability <0.56) risk. We did not refine the estimate of the exposed population in each block due to a lack of additional information, such as the percentage of households with access to treated water or the percentage of people consuming contaminated water, and due to the uncertainty involved in such estimation. Thus, our population estimation could be a slight overestimation. The piped water supply schemes (PWSS) coverage was assessed to determine the status of safe water access in different blocks and analyzed for public health interventions (Integrated Management Information System, 2022).

## 4. Results and discussion

### 4.1 Spatio-temporal pattern of As concentration in groundwater

As concentrations in different blocks were compared using the datasets collected one decade apart (Table S1). The results show that the proportions of As concentrations >10 µg/L are significantly greater in the areas to the east of the Bhagirathi River during the two time periods (Fig. 2). Prior to 2005, 11 out of 26 blocks had a median As concentration <10 µg/L, and most of these blocks are located west of the Bhagirathi River. The pattern in 2015 is similar to that in 2005, which shows that the median As concentration remained <10 µg/L in blocks to the west of the Bhagirathi River. However, the proportion of As concentrations >10 µg/L changed significantly east of the Bhagirathi River over the 10-year period. Most of the As-contaminated blocks east of the Bhagirathi River have shown a decrease in the proportion of As concentrations >10 µg/L (Fig. 3). Such a striking pattern of declining As concentration suggests either flushing of the aquifer or a mixing of high and low As concentrations within the aquifer. However, some exceptions exist, especially in the Hariharpara, Berhampore, Beldanga-II, and Msd-Jiaganj blocks, where the proportion of As concentrations >10 µg/L showed a significant increase. In the Hariharpara block, the proportion of As concentrations >50 µg/L also increased significantly. Marginal increases in the proportion of As concentrations >50 µg/L were noted in the Berhampore and Msd-Jiaganj blocks. In the Lalgola, Bhagawangola-I, Raghunathganj-II, and Suti-II blocks, including the blocks west of the Bhagirathi River, the proportion of As concentrations >10 µg/L in groundwater remained unchanged (Fig. 3).

**Figure 2.**
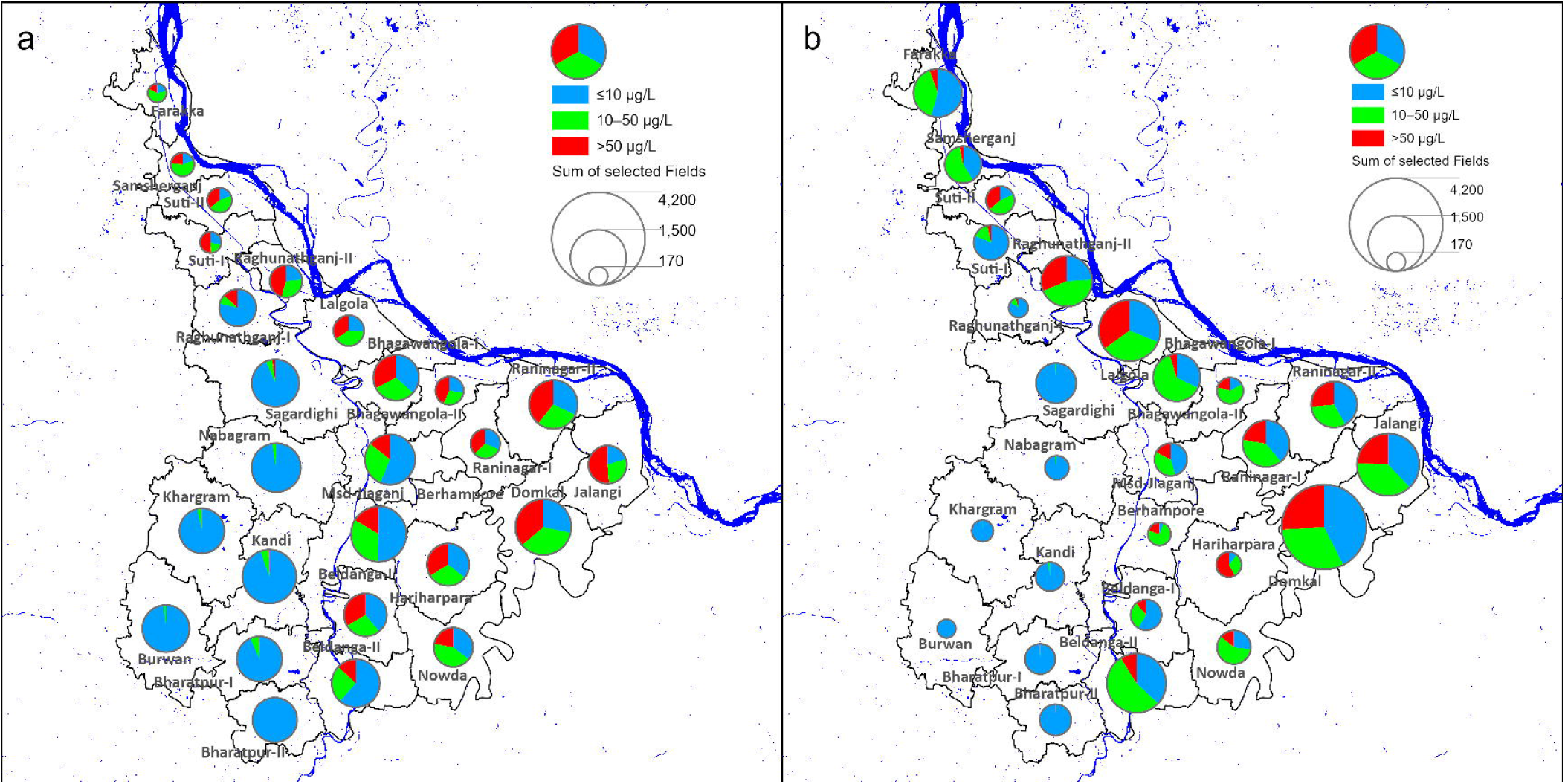
The pie chart shows As in different concentration ranges measured in tube wells: a) prior to 2005 and b) from 2015 and later. The symbol size indicates the total samples in each block.

**Figure 3.**
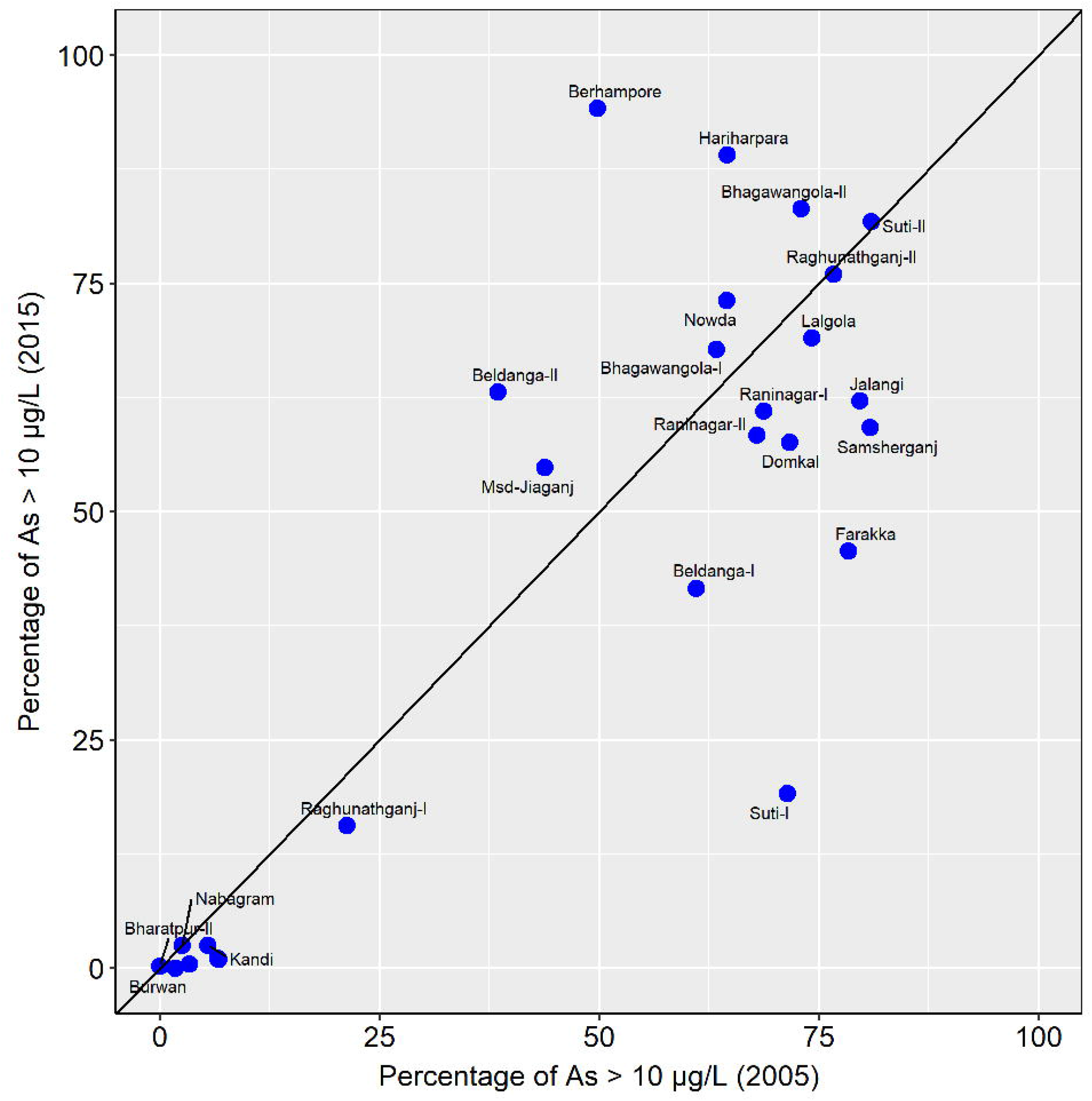
The scatter plot shows the change in the proportion of As concentrations >10 μg/L in different blocks between 2005 and 2015.

To avoid analytical uncertainty when using different subsets of wells to compare As concentrations measured during the two time periods, we further compared the data collected from the Raninagar-II block. In this block, As concentrations in both years were measured with hydride-generation atomic absorption spectroscopy (HG-AAS) in the SOES laboratory of Jadavpur University (Rahman et al., 2005; Das et al., 2021). The results showed that the proportion of As concentrations >10 µg/L decreased from 68% to 58%, suggesting positive steps toward declining population-level exposure to elevated As concentrations. Our analysis further showed that the mean As concentrations in most blocks situated in the eastern part of Murshidabad decreased over the ten years. Only in the Hariharpara block did the mean As concentrations increase significantly (from 80 µg/L to 96 µg/L), along with an increase in the proportions of As concentrations >10 µg/L (from 65% to 89%).

McArthur et al. (2010) reported that As concentrations in groundwater in West Bengal could systematically increase (in 8 out of 20 monitoring wells), decrease (3 out of 20), or remain stable (9 out of 20) in the aquifer over a long time. They argued that their records showed evidence of flushing with low As surface water in shallow aquifers, as well as the contamination of deeper Pleistocene aquifers due to irrigation pumping. Particularly compelling was a documented increase in As concentration during a two-year period when a well was used by the surrounding community, followed by a return to the originally low As levels after pumping ceased (see Fig. 8 in McArthur et al. 2010). Therefore, the periodic monitoring of wells is necessary from a public health perspective, which could further aid in identifying changing patterns of As hotspots.

### 4.2 Modeled probability of As exceedance and model uncertainty

By converting As measurements from 15,335 point locations to 5,326 grid-averaged (250 m) cells, the training and testing data can be uniformly distributed throughout the study area. The uniform distribution of training and testing data is useful from a modeling perspective since the model can be trained to take environmental conditions into account for the entire predicted area. We used a single model in our prediction analysis because the model could be trained for the range of environmental predictor conditions. The density of the training and testing data is adequate for this purpose (Podgorski et al., 2020). We performed 10-fold cross-validation using the training dataset for the final model. The area under the curve (AUC) of the test dataset derived a value of 0.85, which indicates a close-to-perfect model (i.e., a model that performs much better than chance, with a value for pure chance being 0.5) (Fig. S1a). The accuracy of the test dataset is 0.79, and the no information rate is 0.5178. The accuracy is significantly greater than the no information rate (p-value <2.2 × 10^−16^) (Table 2). Cohen’s kappa is 0.5874, which suggests high reliability in our prediction and good model agreement. The model-derived parameters are comparable to several other recent As exceedance prediction studies (Podgorski et al., 2020; Mukherjee et al., 2021). The cutoff value is 0.56, which was used in the development of the hazard probability map (Fig. S1b). The choice of cutoff was made to avoid any bias toward predicting either high or low As concentrations.

**Table 2.** The confusion matrix and statistics of the random forest model after 10-fold cross-validation. The probability cutoff value is 0.56.

| **Predicted class** | **Actual class** |  |
| --- | --- | --- |
|  | 0 | 1 |
| 0 | 596 | 99 |
| 1 | 231 | 671 |
| Accuracy | 0.79 |  |
| Cohen's kappa | 0.59 |  |
| Sensitivity | 0.72 |  |
| Specificity | 0.87 |  |
| Positive predicted value | 0.86 |  |
| Negative predicted value | 0.74 |  |
| Prevalence | 0.52 |  |
| AUC | 0.85 |  |

The model showed that the probability of As concentrations >10 µg/L is mostly greater than 0.6 to the east of the Bhagirathi River, while to the west of the Bhagirathi River, the probability is typically less than 0.3 (Fig. 4a). The highest probability (>0.8) zone follows an N-S longitudinal trend and lies in the interfluvial region of two adjacent river channels (Ganges and Bhagirathi). The identified areas with the highest contamination probabilities include the Lalgola, Bhagawangola-I, Bhagawangola-II, Raninagar-I, Hariharpara, and Nowda blocks. The highest probability zone also contains parts of the Msd-Jiaganj and Berhampore blocks. The modeled probability of As occurrences >10 µg/L in the aquifer highlights the region in which population-level exposure to elevated As concentrations is high and thus requires immediate public health intervention.

**Figure 4.**
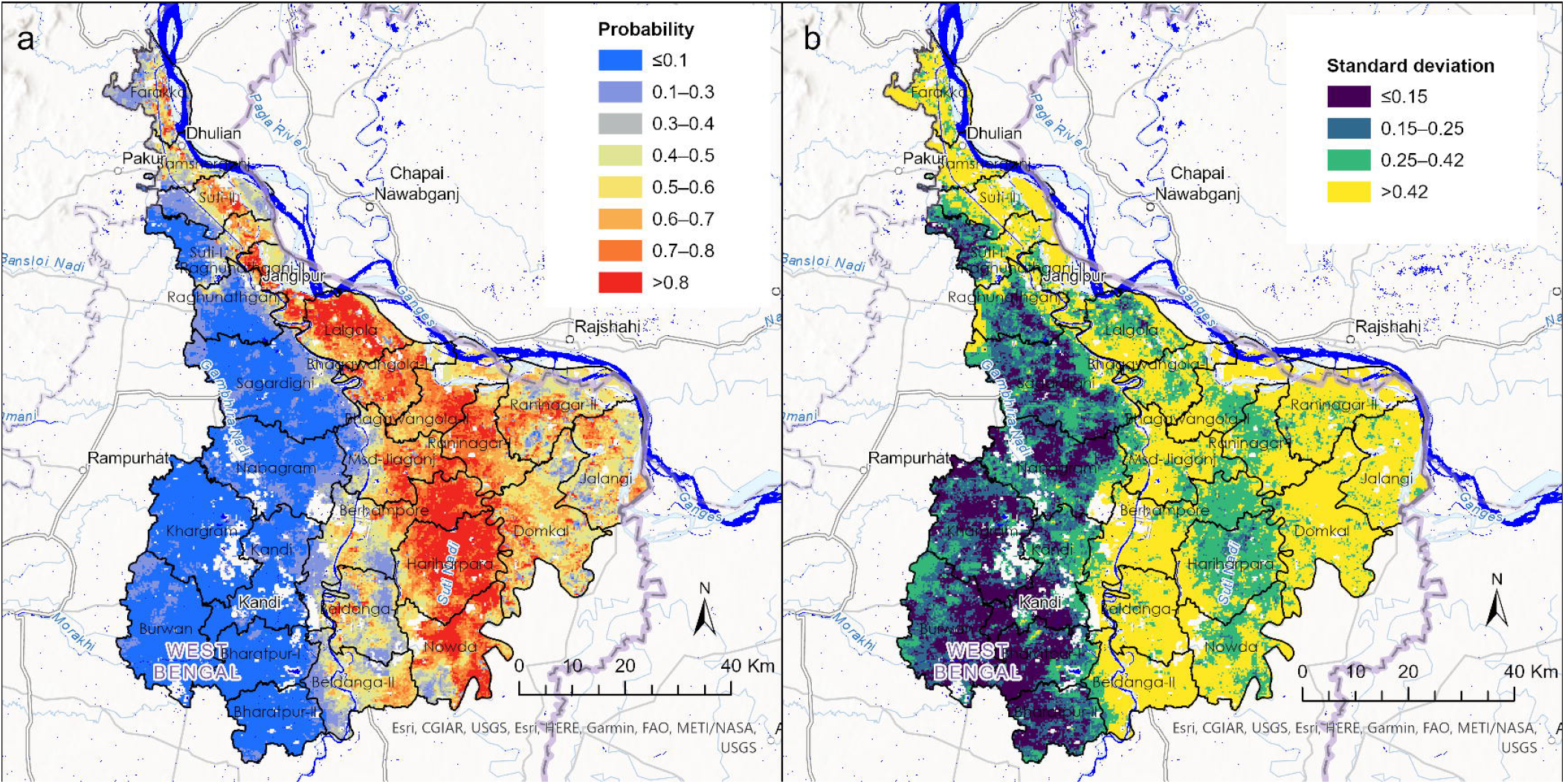
(a) The probability of an As concentration >10 μg/L in groundwater predicted by the random forest model is shown. (b) The standard deviation of the prediction is shown for each grid location.

Model uncertainty was tested using the standard deviation of the predicted class from all trees grown (Fig. 4b). A model with a high and low predicted probability of As concentrations >10 μg/L is generally associated with a low standard deviation. It indicates greater certainty in modeled results because the probabilities derived by the random forest model are the mean values of the outcome of all trees grown during model development (Podgorski et al., 2018). The lowest standard deviation (≤0.15) was observed in much of the area to the west of the Bhagirathi River, which coincided with the areas of lowest probability (<0.3) of As occurrences. Likewise, a lower standard deviation (<0.42) is associated with the highest probability (>0.8). However, the model is less certain (with a standard deviation between 0.42 and 0.50) in the areas near the banks of the Bhagirathi and the Ganges Rivers, coinciding with low to moderate predicted probability. These areas also have a relatively lower sampling density compared to other areas and thus could have affected the overall modeling outcome.

The “area of applicability” of the model was checked by comparing the values of the predictors of the entire study site with those of the training datasets (Meyer and Pebesma, 2021). The calculated dissimilarity index for all predicted pixels within the study area was found to be between 0 and 1: the range considered reliable for model prediction, as suggested by Podgorski et al. (2022).

### 4.3 Role of environmental predictors and their characteristics

The results showed greater importance of that climate variables (temperature, precipitation, potential evapotranspiration, and aridity), fluvisols, topsoil sand, and topsoil clay were more important in the model prediction than other variables (Fig. 5). Topsoil and subsoil cation exchange capacity, together with subsoil sand, subsoil clay, topsoil silt, and subsoil silt, also showed strong importance in predicting high and low As concentrations. The least important variables were subsoil bulk density, topsoil bulk density, and gleysols. The scores of mean decrease in Gini and accuracy consistently showed high importance for the temperature, fluvisols, potential evapotranspiration, and aridity variables, in contrast to low importance for the subsoil bulk density and top bulk density variables. All of the predictors showed beneficial effects on the model since none of the predictors had a negative importance value (Fig. 5).

**Figure 5.**
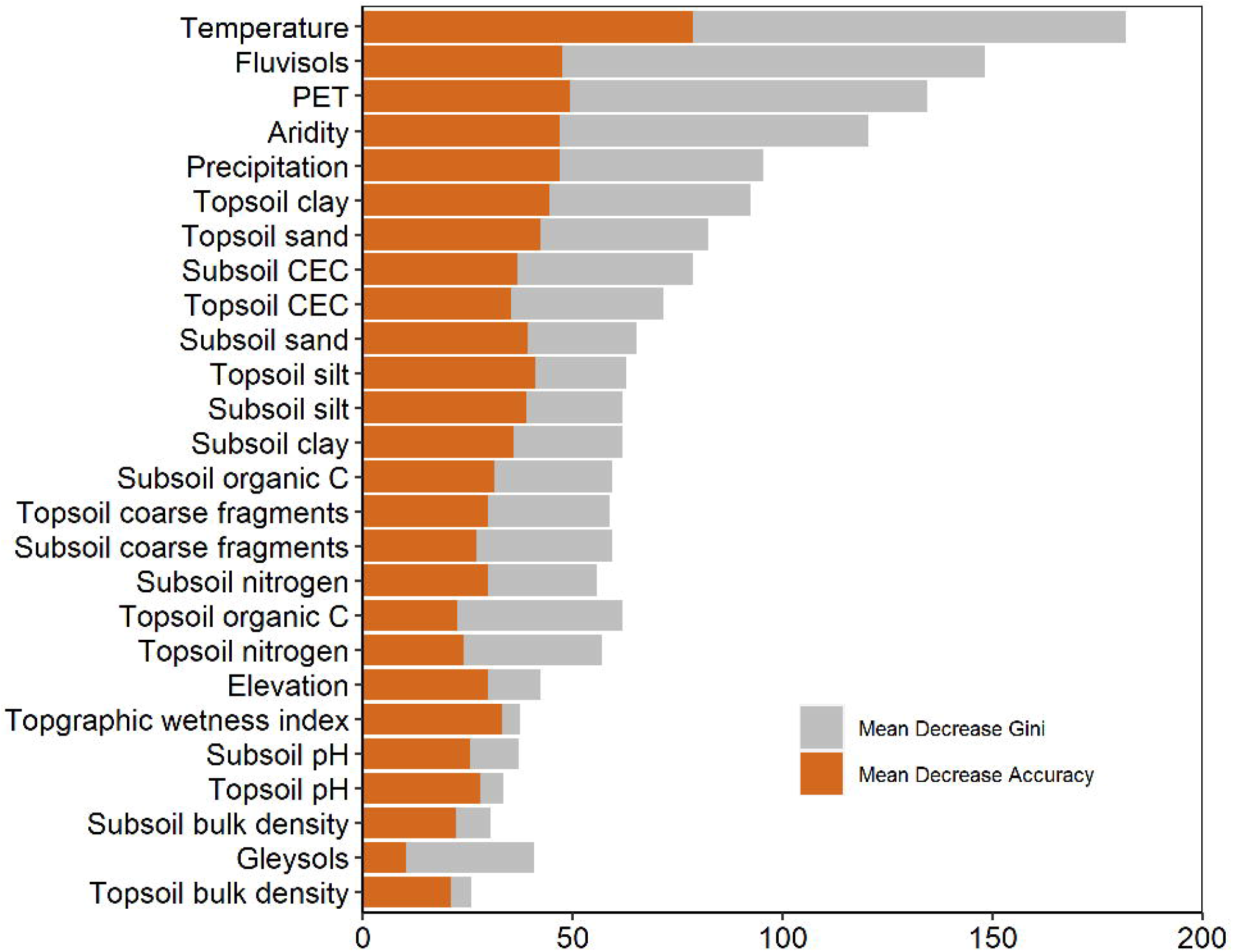
The relative importance of predictor variables is shown based on the values of mean decrease in Gini and mean decrease in accuracy. The greater relative importance of the predictor variables is indicated by the larger values.

Topsoil clay, subsoil clay, topsoil organic carbon, topsoil sand, subsoil sand, topsoil cation exchange capacity, subsoil cation exchange capacity, topsoil bulk density, topsoil coarse fragments, subsoil coarse fragments, temperature, potential evapotranspiration, elevation, fluvisols, and gleysols, showed strong relationships with the proportions of As concentrations >10 µg/L. The R values were >0.7 (either positive or negative) and p-values <0.05 in all of these cases (Fig. 6).

**Figure 6.**
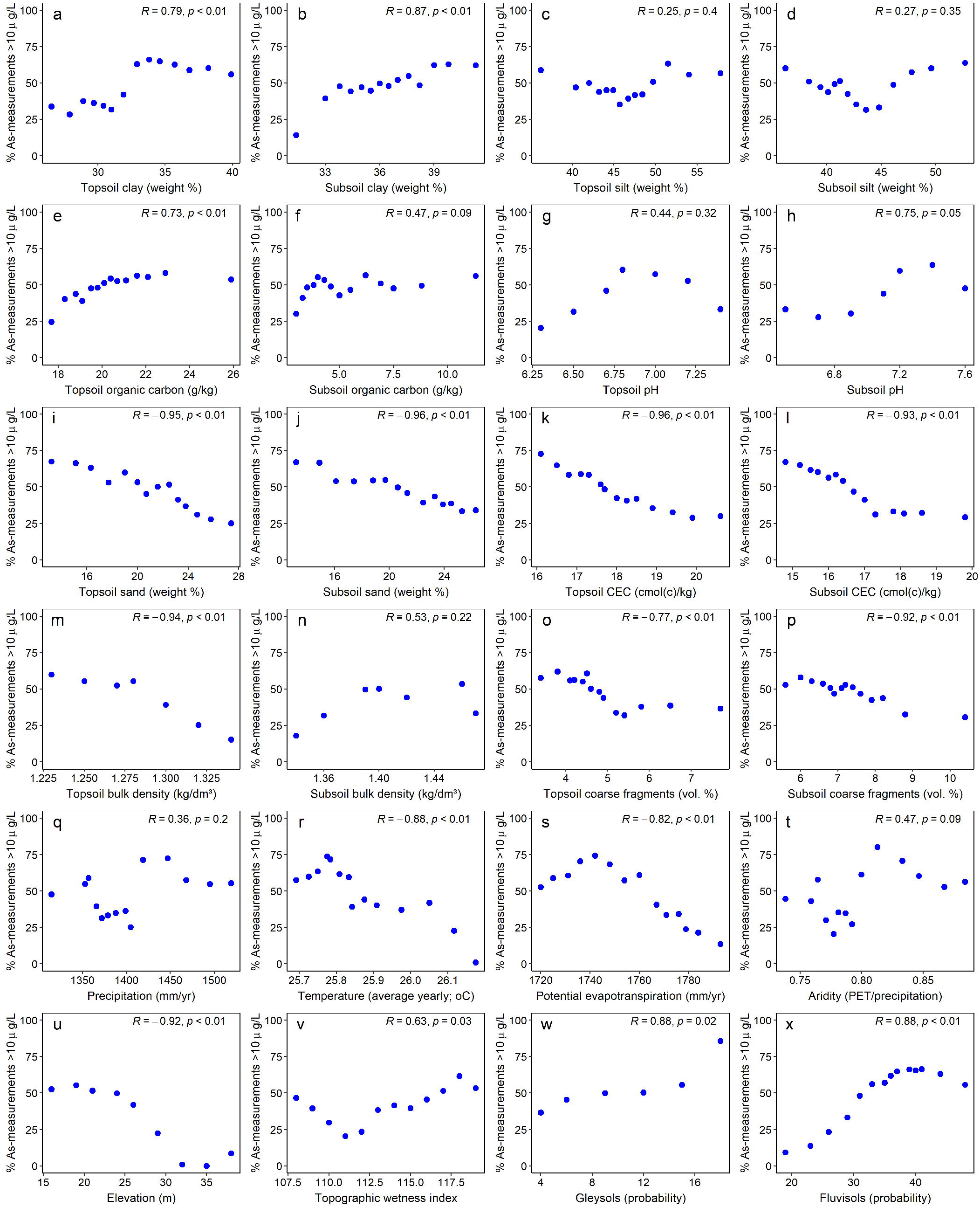
The correlation between the predictor variable (**a–x**) and the proportion of As concentrations >10 µg/L in 14 equally sized bins. Statistical significance is represented by p-values (95% confidence level) and Pearson correlation coefficients (R).

High importance to fluvisols probability, including a strong positive correlation with the proportion of As concentrations >10 µg/L, indicates that the occurrence of high As concentrations is limited to floodplains. Fluvisols are very young soils, typically associated with alluvial deposits, which are more enriched with As than soils at a distance from the river flows (Barać et al., 2015). Likewise, a positive correlation between gleysols’ probability and As concentration indicates reducing environments favoring As release in the aquifer (Podgorski et al., 2020). Topsoil clay is also highly important in our model prediction. Its strong positive correlation with the proportion of As concentrations >10 µg/L supports the occurrence of elevated As concentrations beneath floodplains, where clay plugs could provide the source of labile organic carbon and help make favorable conditions for As to be released from sediments into groundwater (Nath et al., 2005; Donselaar et al., 2017). The association between topsoil organic carbon and the occurrences of As groundwater in alluvial settings is favored for redox-induced mobilization of As in groundwater. Surficial capping by clayey materials would favor the development of reducing conditions and releasing As into groundwater (Nath et al., 2010; Choudhury et al., 2018). Like many other studies, our model also showed the strong control of elevation. Typically, high levels of As are present in low-lying, flat topographical areas, where biogeochemical conditions favor the development of extended water-rock interactions due to slugging groundwater movement and the development of strong reducing conditions. Conversely, higher surface and sub-surface sand fractions indicate a groundwater recharge zone and potentially an unfavorable location for As release due to the abundant presence of oxygenated groundwater recharge (Nath et al., 2009). A negative correlation between topsoil and subsoil sand with As concentrations support such processes.

The role of climate variables on As accumulation in groundwater has been described previously (Podgorski and Berg, 2020). The combination of high precipitation, low temperature, and low potential evapotranspiration favors waterlogged conditions and increases the likelihood of As release under reducing conditions.

### 4.4 Arsenic hazard probabilistic zones and population exposed

A probability cutoff value of 0.56 was used to compute at-risk probabilistic zones and the population exposed in the study area (Table S2 and Fig. 7). According to the hazard probability map, 30% of the study area is at moderate and high risk of exposure to As concentrations >10 µg/L. The high-risk areas are primarily located in the interfluvial regions of two adjacent rivers, the Bhagirathi and the Ganges. The low-risk areas are primarily located to the west of the Bhagirathi River, yet isolated high-risk pockets are located in these low-risk areas. Such a difference in groundwater As probabilistic zones on either side of the Bhagirathi River has been attributed to the difference in aquifer sediment types (Datta et al., 2011). A previous study documented that the aquifer to the west of the Bhagirathi River is dominated by Pleistocene orange sands, while to the east, gray Holocene sands dominate (Chatterjeee et al., 2020). Orange Pleistocene sands have much greater adsorption capacity than gray Holocene sands and thus control aquifer As distribution (van Geen et al., 2013). Most of the areas near the two adjacent rivers are either low- or moderate-risk. These low to moderate probabilistic zones for As near the river could be attributed to the reversal of the groundwater flow pattern since intense agricultural pumping could make the discharging river a losing stream and provide oxygenated recharge to the aquifer (Stahl et al., 2016).

**Figure 7.**
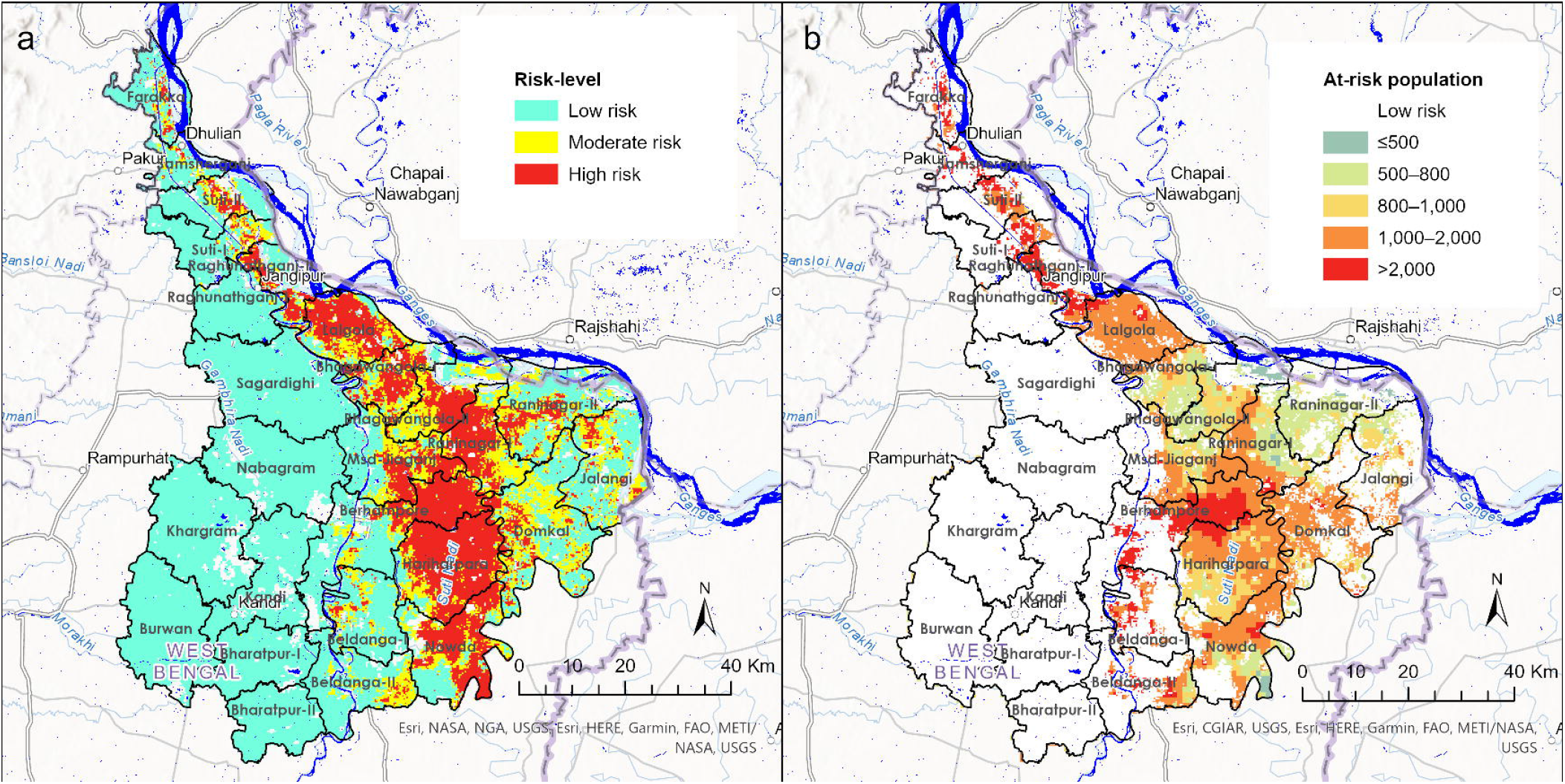
(a) A probabilistic As hazard map was prepared based on the probability values derived from the random forest model (high risk: ≥0.7, moderate risk: 0.56–0.7, and low risk: <0.56). (b) The population density (per km^2^) in high- and moderate-risk areas is shown.

Based on the cutoff probability of 0.56, a total of approximately 2,758,547 people (one-third of the total population) are at risk of being exposed to As concentrations >10 µg/L. The calculated at-risk populations could be refined further based on knowledge of the exact number of houses connected to government-owned treated piped water connections. Moreover, As exposure through the food chain cannot be ruled out since most agricultural land uses groundwater for irrigation (Haque, 2015). The proportion of at-risk (moderate- and high-risk) populations is documented block-wise (Fig. S2).

This study applies a unique method—a machine learning technique—to compute at-risk populations at the block (or sub-district) level in West Bengal, India. The machine learning computation of at-risk populations is significantly less than the computation of exposed populations by Rahman et al. (2005). Most exposed populations live in five blocks: Berhampore, Hariharpara, Lalgola, Nowda, and Domkal (Fig. 8). Most of these people require alternative drinking water sources, as only 30 to 44% of the households in these blocks are connected to safe water through a piped network (Fig. 8). In addition, many people are at risk in the Domkal, Msd-Jiaganj, and Raninagar-I blocks and also require alternative drinking water sources.

**Figure 8.**
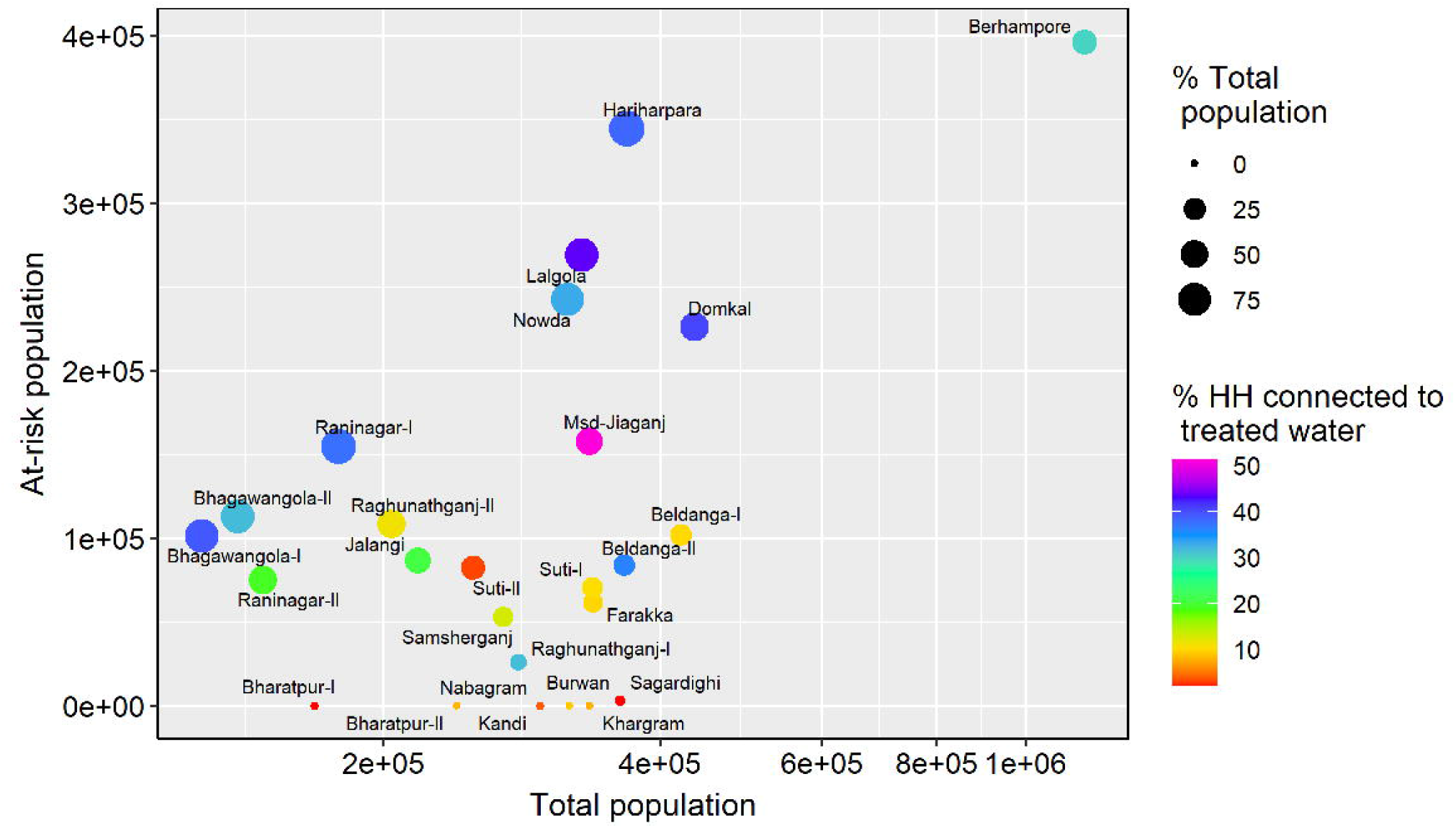
The scatter plot shows the at-risk population against the total population in different blocks. The symbol size represents the percentage of the total population. The symbol color represents the percentage of households (HH) connected to public water supply schemes (PWSS) providing treated water.

## 5. Public health considerations and mitigation measures

The probabilistic risk map we generated for As exceedance in groundwater above the WHO provisional guideline value of 10 µg/L will help policymakers manage the aquifer sustainably, since agricultural groundwater use is widespread in the study area (Haque, 2015). In most of the highly affected blocks, groundwater irrigation covers more than 80% of the area (Haque, 2015). Chatterjeee et al. (2020) observed a significant declining trend in groundwater levels in the study area. The decline in groundwater level, together with intense irrigation pumping, could facilitate instability of the aquifer’s geochemical conditions by altering groundwater flow dynamics and aquifer mixing. Therefore, regular well-testing and subsequent well-switching could effectively lower population-level exposure to elevated As concentrations in high-risk areas lacking a treated, household piped water supply. Due to strong spatial heterogeneity and the absence of permanent safe water solutions, researchers have previously proposed well-switching to reduce population-level exposure to elevated As concentrations (van Geen et al., 2002). One of the well-switching strategies that could be adopted by the affected households is to switch their drinking and cooking water sources from high-As to the nearest low-As shallow wells. This strategy has gained considerable popularity relative to other proposed options in Bangladesh, with nearly 29% of the initially exposed population switching their drinking water sources to the nearest low-As shallow wells (Ahmed et al., 2006). Well-switching could be effective in high-risk areas, as a previous study showed that As occurrence in groundwater is spatially heterogenous (Mukherjee et al., 2010). Predictive modeling at habitation scale thus becomes advantageous in the short term and provides evidence for the necessity of well-switching options. However, the transfer of As from groundwater to the food chain remains a threat to human health since there is increased groundwater-fed irrigation in Murshidabad (Haque, 2015; Das et al., 2021).

The analysis of PWSS coverage revealed that house connections remain below 40% of the total households in five high-risk blocks. This finding suggests a heightened risk of population-level exposure to high As concentrations in many rural households due to the lack of access to safe water. Though public standposts provide tap water connections available for use in many unconnected rural households in high-risk blocks, a report suggests public reluctance to travel long distances to get cleaner water (Nahar et al., 2008). The Lalgola, Bhagawangola-I, and Bhagawangola-II blocks are among the worst regions in terms of contamination, as both PWSS house connections and public standposts are lacking (Fig. S3). These blocks lack access to safe drinking water through private or public tapwater connections; about 22–27% of households have not yet been served with PWSS water. Therefore, it is important from a public health perspective to formulate plans to generate awareness among rural inhabitants, especially those in high-risk blocks, to use community piped water for their drinking water needs or to switch from high-As wells to low-As wells.

## 6. Conclusion

This application of geospatial machine learning techniques provides the first detailed picture of the block (sub-district)-level distribution of elevated As in the Murshidabad district of West Bengal, India. The comparison of As concentrations measured in well water after a decade highlights possible shifts in hotspot locations, with a greater proportion of elevated As (>10 µg/L) in the study area. A possible reason for such shifts in hotspot location could be the intensive groundwater pumping for agricultural production in the region between the two adjacent river channels. The spatio-temporal pattern of As concentrations in different blocks indicates the likely impact of groundwater pumping on aquifer As stability. Over a decade, the threat of exposure to elevated As concentrations has changed spatially toward high population density villages.

The risk map we generated based on recent As measurements could provide up-to-date information to local inhabitants on the potential danger of elevated As concentrations in their groundwater and could inform policymakers in formulating an action plan for mitigation. The ternary risk map could be used to prioritize locations for intervention and serve the community through drinking water network wells to facilitate safe water access. A targeted well-testing campaign could encourage well-switching (from high-As to low-As wells) to avoid drinking contaminated water.

Groundwater As has considerable spatial heterogeneity, which should be considered in future predictive models. Iron and manganese are the two major constituents of groundwater and correlate strongly with As, and future models should include these two predictors to improve the model.

## Data Availability

Available upon reasonable request to corresponding author.

## Acknowledgments

This study was funded by NASA grant number 80NSSC20K1718.

## Conflict of interest

The authors declare no conflicts of interest in this study.

## Notes

### Competing Interest Statement

The authors have declared no competing interest.

