## Supplemental figures and tables for "A geospatial machine learning prediction of arsenic distribution in the groundwater of Murshidabad district, West Bengal, India: spatio-temporal pattern and human health risk"

**Contents of this file**

Figures S1 to S3

Tables S1 to S2

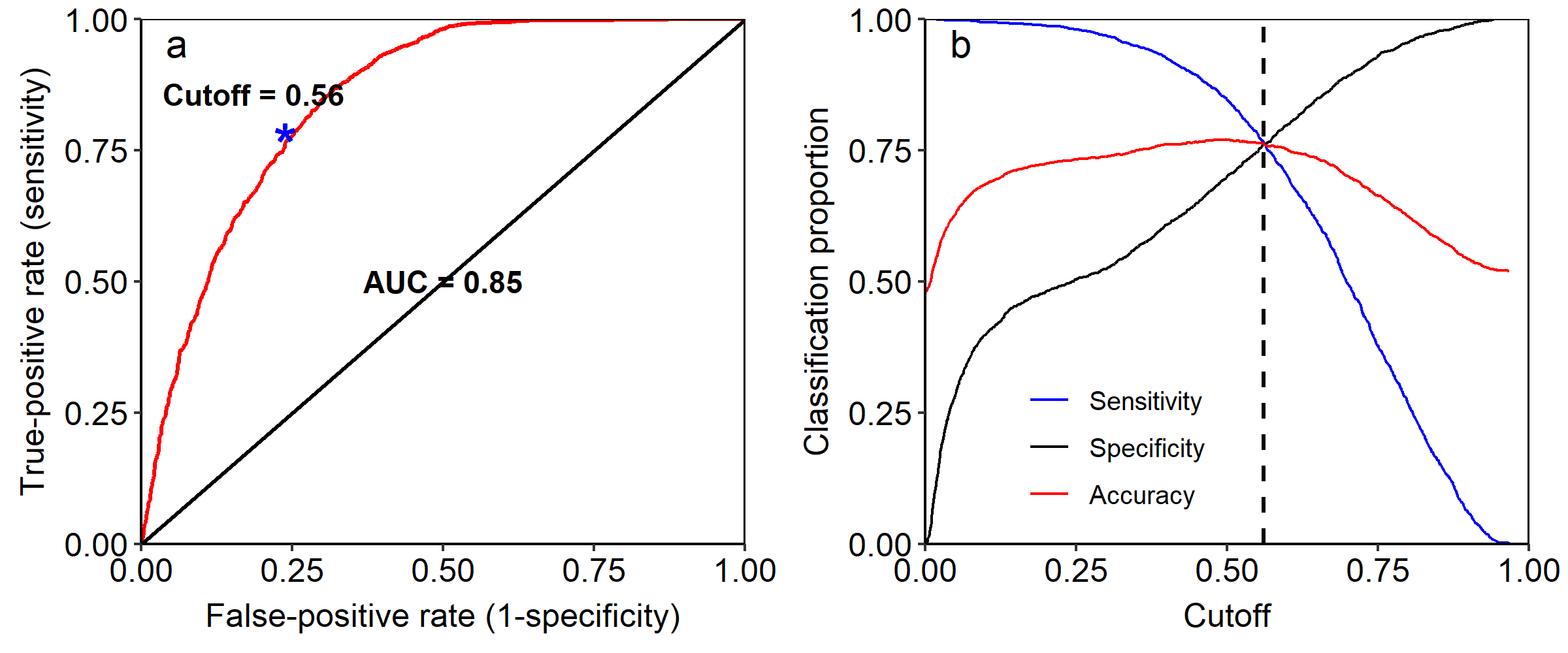

Fig. S1. (a) The ROC curve with an AUC of 0.85. The AUC value is significantly greater than 0.5, indicating the strength of the model. (b) Sensitivity, specificity, and accuracy values were plotted against the different cutoff values. The best cutoff (0.56) was chosen when sensitivity and specificity intersect in predicting low and high As concentrations.

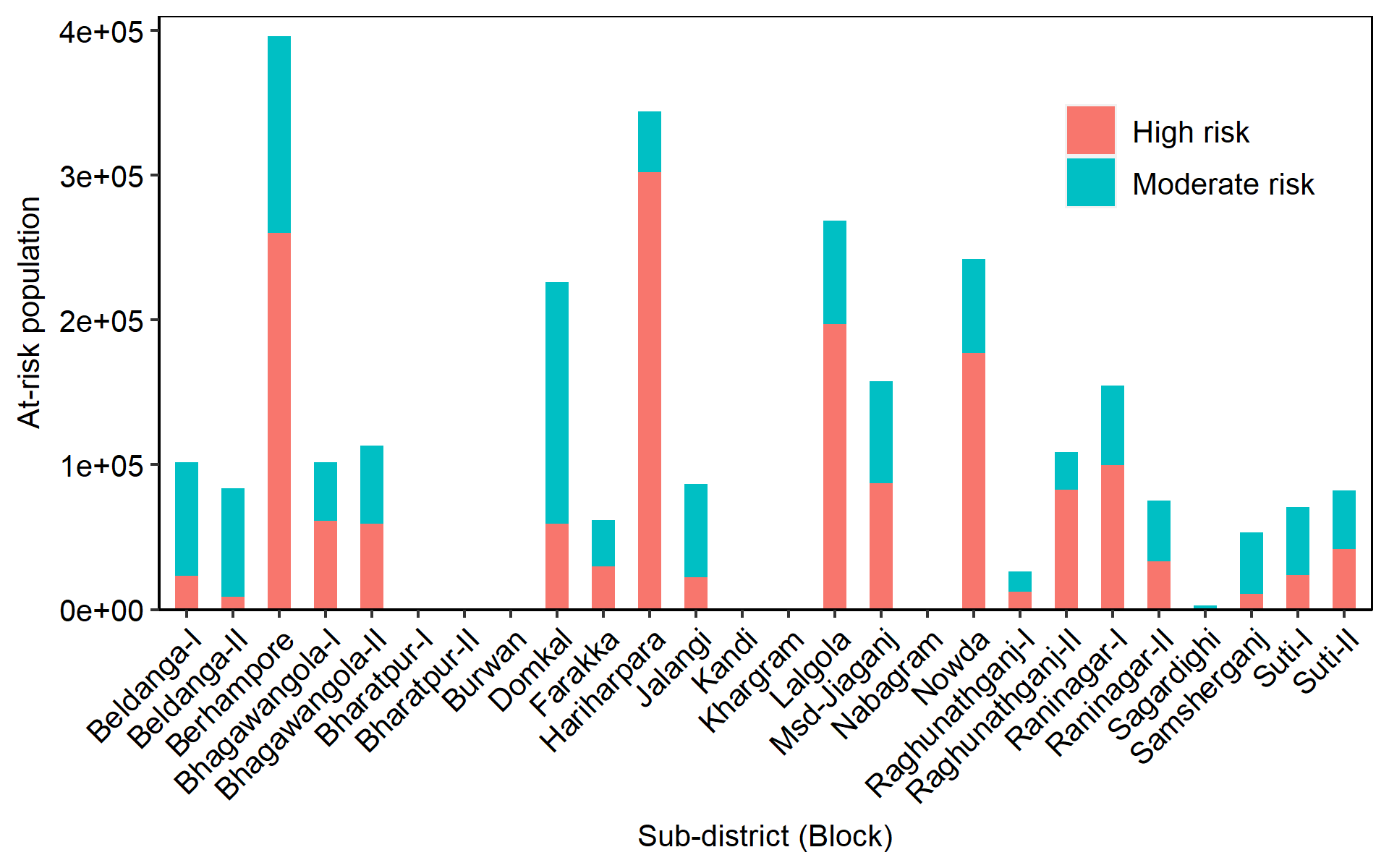

Fig. S2. At-risk population in high and moderate risk areas in twenty-six blocks (sub-districts) of the study area.

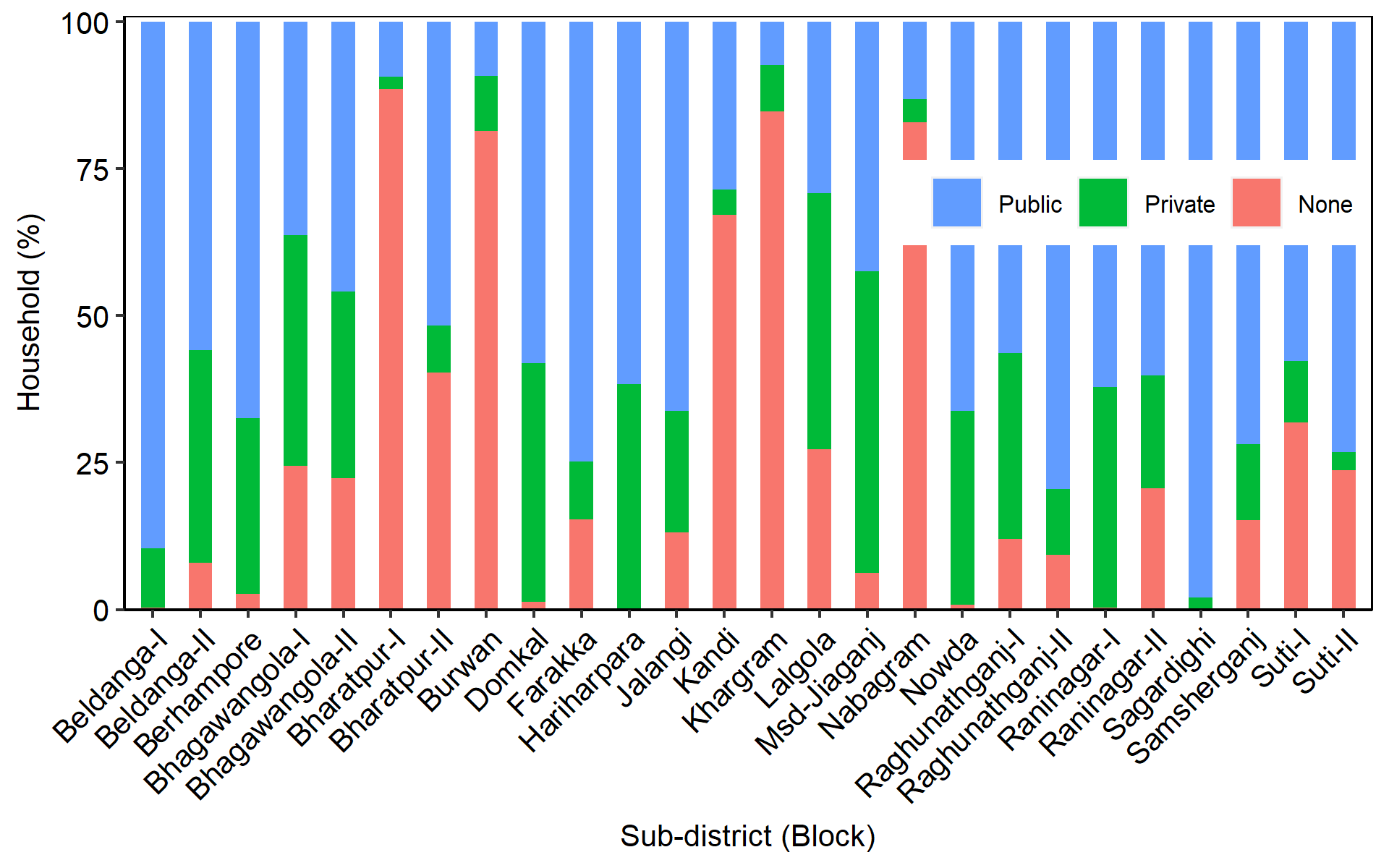

Fig. S3. Percent household across three categories: access to treated water through 'public' connections, access to treated water through 'private' connections, and no connectivity ('none') to treated water in twenty-six blocks (sub-districts) of the study area.

Table S1. Summary statistics of As concentrations in groundwater measured before 2005 and after 2015.

| **Blocks** | **Year 2005** | | | | | | **Year 2015** | | | | | |
| --- | --- | --- | --- | --- | --- | --- | --- | --- | --- | --- | --- | --- |
|  | *Sample* | *As*  *>10 µg/L* | *As*  *>50 µg/L* | *As_mean_* | *As_median_* | *As_std_dev_* | *Sample* | *As*  *>10 µg/L* | *As*  *>50 µg/L* | *As_mean_* | *As_median_* | *As_std_dev_* |
| Beldanga-I | 1193 | 729 | 401 | 77 | 21 | 149 | 459 | 191 | 53 | 52 | 9 | 316 |
| Beldanga-II | 948 | 365 | 124 | 23 | 3 | 42 | 1701 | 1073 | 150 | 21 | 11 | 31 |
| Berhampore | 1545 | 770 | 256 | 29 | 9 | 54 | 258 | 243 | 52 | 37 | 27 | 42 |
| Bhagawangola-I | 1436 | 910 | 469 | 64 | 24 | 106 | 1087 | 737 | 53 | 18 | 14 | 21 |
| Bhagawangola-II | 752 | 549 | 324 | 103 | 38 | 175 | 339 | 282 | 71 | 34 | 30 | 32 |
| Bharatpur-I | 552 | 37 | 1 | 4.4 | 3 | 6.1 | 441 | 4 | 1 | 4.4 | 2 | 34 |
| Bharatpur-II | 502 | 0 | 0 | 3 | 3 | 0 | 477 | 1 | 1 | 5.3 | 5 | 4.2 |
| Burwan | 571 | 10 | 2 | 3.4 | 3 | 3.9 | 169 | 0 | 0 | 2.5 | 2 | 1.6 |
| Domkal | 2796 | 2005 | 1014 | 80 | 30 | 127 | 3491 | 2009 | 907 | 41 | 15 | 70 |
| Farakka | 407 | 319 | 72 | 29 | 24 | 23 | 1135 | 518 | 62 | 16 | 9 | 19 |
| Hariharpara | 1273 | 822 | 431 | 80 | 21 | 135 | 312 | 278 | 180 | 96 | 62 | 112 |
| Jalangi | 1822 | 1452 | 934 | 155 | 53 | 245 | 1888 | 1173 | 460 | 44 | 15 | 78 |
| Kandi | 770 | 42 | 5 | 4.3 | 3 | 5.9 | 401 | 10 | 0 | 1.2 | 0 | 3.6 |
| Khargram | 528 | 18 | 2 | 3.6 | 3 | 4.0 | 229 | 1 | 0 | 3.6 | 4 | 1.4 |
| Lalgola | 919 | 682 | 312 | 59 | 33 | 99 | 1829 | 1263 | 640 | 48 | 27 | 58 |
| Msd-Jiaganj | 1139 | 499 | 163 | 23 | 7 | 35 | 518 | 284 | 91 | 26 | 10 | 38 |
| Nabagram | 624 | 16 | 0 | 3.5 | 3 | 2.5 | 283 | 7 | 0 | 3.1 | 3 | 2.1 |
| Nowda | 1092 | 705 | 241 | 49 | 20 | 129 | 550 | 402 | 77 | 31 | 18 | 52 |
| Raghunathganj-I | 441 | 94 | 59 | 57 | 3 | 322 | 186 | 29 | 8 | 8.7 | 4 | 15 |
| Raghunathganj-II | 1141 | 875 | 522 | 62 | 43 | 79 | 1254 | 953 | 391 | 43 | 31 | 47 |
| Raninagar-I | 710 | 488 | 262 | 56 | 25 | 77 | 1052 | 642 | 236 | 39 | 15 | 64 |
| Raninagar-II | 1871 | 1272 | 735 | 105 | 27 | 160 | 1023 | 597 | 275 | 74 | 14 | 147 |
| Sagardighi | 606 | 40 | 13 | 6.2 | 3 | 25 | 778 | 8 | 1 | 3.9 | 3 | 3.4 |
| Samsherganj | 738 | 597 | 172 | 43 | 26 | 52 | 654 | 387 | 21 | 12 | 10 | 17 |
| Suti-I | 406 | 290 | 207 | 79 | 53 | 95 | 592 | 113 | 22 | 8.3 | 2 | 21 |
| Suti-II | 821 | 665 | 295 | 82 | 32 | 151 | 394 | 322 | 143 | 54 | 30 | 66 |

Table S2. Summary statistics of risk population, risk area, PWSS connectivity and groundwater-fed irrigation.

| **Blocks** | **Total**  **population** | **Risk**  **population^a^** | **High-risk**  **Population^b^** | **Total**  **Area**  **(Km^2^)** | **Risk**  **area (%)^a^** | **High-risk**  **area (%)^b^** | **PWSS_public_**  **(% HH)** | **PWSS_private_**  **(% HH)** | **PWSS_None_**  **(% HH)** | **GW irrigation**  **(% area,**  **1994-95)** | **GW irrigation**  **(% area,**  **2010-11)** |
| --- | --- | --- | --- | --- | --- | --- | --- | --- | --- | --- | --- |
| Beldanga-I | 421336 | 101719 | 23367 | 182 | 21 | 4.9 | 89.6 | 10.06 | 0.34 | 71.71 | 73.94 |
| Beldanga-II | 365900 | 83862 | 8821 | 197 | 22 | 2.9 | 55.9 | 36.15 | 7.95 | 79.17 | 81.52 |
| Berhampore | 1158451 | 396196 | 260383 | 320 | 43 | 32 | 67.39 | 30 | 2.61 | 76.5 | 77.68 |
| Bhagawangola-I | 126799 | 101496 | 61237 | 127 | 72 | 43 | 36.22 | 39.37 | 24.41 | 90.64 | 96.41 |
| Bhagawangola-II | 138561 | 113063 | 59019 | 186 | 62 | 28 | 45.82 | 31.77 | 22.41 | 93.71 | 95.73 |
| Bharatpur-I | 167998 | 142 | 0 | 172 | 0.07 | 0 | 9.33 | 2.03 | 88.64 | 71.39 | 28.51 |
| Bharatpur-II | 240188 | 0 | 0 | 171 | 0 | 0 | 51.6 | 8.09 | 40.3 | 60.11 | 28.31 |
| Burwan | 318960 | 0 | 0 | 297 | 0 | 0 | 9.21 | 9.31 | 81.48 | 71.1 | 72.57 |
| Domkal | 436096 | 226286 | 59322 | 321 | 51 | 13 | 58.02 | 40.64 | 1.34 | 81.74 | 85.35 |
| Farakka | 338013 | 61649 | 29785 | 144 | 13 | 6.3 | 74.75 | 9.91 | 15.34 | 26.42 | 36.59 |
| Hariharpara | 368107 | 344369 | 302005 | 262 | 91 | 78 | 61.67 | 38.33 | 0 | 73.03 | 81.97 |
| Jalangi | 217992 | 86687 | 22090 | 228 | 37 | 9.6 | 66.24 | 20.63 | 13.14 | 90.6 | 89.87 |
| Kandi | 296630 | 0 | 0 | 255 | 0 | 0 | 28.49 | 4.34 | 67.17 | 56.73 | 55.71 |
| Khargram | 335083 | 0 | 0 | 318 | 0 | 0 | 7.35 | 7.9 | 84.75 | 43.33 | 32.32 |
| Lalgola | 328503 | 268909 | 197052 | 182 | 77 | 56 | 29.08 | 43.61 | 27.31 | 83.8 | 80.38 |
| Msd-Jiaganj | 334746 | 157945 | 87202 | 201 | 54 | 30 | 42.48 | 51.29 | 6.23 | 53.44 | 44.36 |
| Nabagram | 295799 | 167 | 82 | 296 | 0.04 | 0.02 | 13.12 | 3.92 | 82.95 | 54.85 | 59.32 |
| Nowda | 317025 | 242229 | 177114 | 248 | 69 | 49 | 66.17 | 33.06 | 0.77 | 54.4 | 72.11 |
| Raghunathganj-I | 280551 | 26058 | 12090 | 158 | 5.0 | 2.1 | 56.3 | 31.68 | 12.01 | 54.6 | 40.41 |
| Raghunathganj-II | 203820 | 108545 | 82863 | 108 | 33 | 25 | 79.46 | 11.23 | 9.3 | 48.28 | 72.34 |
| Raninagar-I | 178404 | 154937 | 99724 | 147 | 85 | 51 | 62.1 | 37.58 | 0.32 | 79.59 | 89.12 |
| Raninagar-II | 147696 | 75230 | 33220 | 243 | 39 | 16 | 60.16 | 19.25 | 20.59 | 96.52 | 97.32 |
| Sagardighi | 361418 | 2972 | 779 | 356 | 0.56 | 0.12 | 97.97 | 2.03 | 0 | 67.4 | 73.06 |
| Samsherganj | 270161 | 53032 | 10700 | 84 | 14 | 2.6 | 71.9 | 12.95 | 15.15 | 82.09 | 71.56 |
| Suti-I | 337419 | 70710 | 23845 | 162 | 17 | 5.3 | 57.64 | 10.51 | 31.85 | 35.71 | 51.14 |
| Suti-II | 250375 | 82345 | 41933 | 138 | 20 | 10 | 73.16 | 3.13 | 23.71 | 64.09 | 66.26 |

Note: ^a^based on probability ≥0.56; ^b^based on probability ≥0.70; and HH = households.
